# The COVID-19 Infodemic: The complex task of elevating signal and eliminating noise

**DOI:** 10.1101/2021.01.19.21249936

**Authors:** Tejas Desai, Arvind Conjeevaram

## Abstract

In Situation Report #3 and 39 days before declaring COVID-19 a pandemic, the WHO declared a -19 infodemic. The volume of coronavirus tweets was far too great for one to find accurate or reliable information. Healthcare workers were flooded with which drowned the of valuable COVID-19 information. To combat the infodemic, physicians created healthcare-specific micro-communities to share scientific information with other providers. We analyzed the content of eight physician-created communities and categorized each message in one of five domains. We coded 1) an application programming interface to download tweets and their metadata in JavaScript Object Notation and 2) a reading algorithm using visual basic application in Excel to categorize the content. We superimposed the publication date of each tweet into a timeline of key pandemic events. Finally, we created NephTwitterArchive.com to help healthcare workers find COVID-19-related signal tweets when treating patients. We collected 21071 tweets from the eight hashtags studied. Only 9051 tweets were considered signal: tweets categorized into both a domain and subdomain. There was a trend towards fewer signal tweets as the pandemic progressed, with a daily median of 22% (IQR 0-42%. The most popular subdomain in Prevention was PPE (2448 signal tweets). In Therapeutics, Hydroxychloroquine/chloroquine wwo Azithromycin and Mechanical Ventilation were the most popular subdomains. During the active Infodemic phase (Days 0 to 49), a total of 2021 searches were completed in NephTwitterArchive.com, which was a 26% increase from the same time period before the pandemic was declared (Days −50 to −1). The COVID-19 Infodemic indicates that future endeavors must be undertaken to eliminate noise and elevate signal in all aspects of scientific discourse on Twitter. In the absence of any algorithm-based strategy, healthcare providers will be left with the nearly impossible task of manually finding high-quality tweets from amongst a tidal wave of noise.

## Introduction

Medical professionals are increasingly using Twitter to share scientific information with each other (1-3). Healthcare providers in many medical specialties are using Twitter to broadcast cases, conduct journal clubs, and share scientific information presented at their respective meetings (1-5). However, not all of this information is accurate, credible, and/or supported by scientific evidence (noise tweets) (6-7). Physician-learners have a difficult time identifying highly educational tweets (signal tweets) because the quality of scientific information created and shared through Twitter is variable. This difficulty worsens every year as the total number of tweets authored or retweeted by physicians increases (8). To our knowledge, there has not been a concerted effort to help physicians eliminate noise and elevate signal tweets.

Perhaps with great foresight, the World Health Organization (WHO) identified the problem of finding signal tweets about the COVID-19 pandemic. Nearly thirty-nine days prior to declaring COVID-19 a pandemic (2 February 2020), the WHO declared a “COVID-19 Infodemic” (9). WHO leaders predicted that the volume of COVID-19-related tweets would be so great that high quality tweets about the disease/pandemic would be buried under an avalanche of lower/poor quality ones (noise). To combat the COVID-19 Infodemic, the WHO committed itself to tweeting the most accurate and evidence-based information about COVID-19 through its official Twitter accounts (9). The Organization did not commit to relaying high-quality information from sources other than itself.

This commitment, while admirable, neglected the physician social media community that has, for years, shared credible and evidence-based scientific information through Twitter (1-5). Beginning on the day of the WHO declared COVID-19 a pandemic (Day 0; 11 March 2020) we undertook an initiative to a) identify high quality tweets regarding COVID-19 from various physician communities, b) categorize these tweets, and c) archive the most informative of these tweets for future searches. Our objective was to eliminate noise and elevate signal tweets related to COVID-19 and provide easy access to the most educational tweets for medical professionals who were searching for information.

## Methods

We identified COVID-19-related tweets from Days 0-199 (11 March to 27 September 2020) from eight physician-specific hashtags established by various members of the physician social media community (10). These hashtags were: #, #4rheum, #4MD, #4MDs, #2, #4MD, #4MDs, and #19Kidney. While physicians from Nephrology, Rheumatology, and Internal Medicine created these hashtags, anyone could read and post within them. The hashtags were created on Days 0 or 1 of the pandemic. They were established to share medical information about COVID-19 with the scientific community but their use was unrestricted and unregulated. We used an application programming interface coded in JavaScript to download the tweets (and its associated metadata) every hour during the study period. All tweets were translated into English using Google Translate. We stored the translated tweets in Microsoft Excel and categorized each into one of five domains and one of 84 subdomains. The five domains were: Diagnostics, Pathophysiology, Prevention, Symptoms, and Therapeutics. The 84 subdomains are listed in Table 1.

**Table 1.**

| **Domain** | **Subdomain** |
| --- | --- |
| *Diagnostic* | testing, swab, test, kit, tracing, Crispr |
| *Pathophysiology* | IL6, ACE, angiotensin, interferon, autophagy, Ly6e protein, antiphospholipid antibody, cytokine, clot, sigma, TLR7 |
| *Prevention* | ppe, mask, n95, respirator, sanitizer, wash hands, 6 feet, six feet, two meters, 2 meters, social distancing, immunity, lockdown, virtual |
| *Symptoms* | dexamethasone, pamrevlumab, Bruton tyrosine kinase, temperature, fever, febrile, anosmia, hyposmia, cough, ST segment, guillain-barre, coagulopathy, subclinical, shortness of breath, sneeze |
| *Therapeutics* | famotidine, llama,vaccine, remdesivir, hydroxychloroquine, azithromycin, bcg, tocilizumab, acetemra, tofacitinib, baracitinib, ventilator, intubate, prone, extracorporeal, hemoperfusion, activated protein c, zinc, ivermectin, favipiravir, colchicine, COLCORONA, sevoflurane, soluble Ace2, lopinavir, ritonavir, PREP, baloxavir, stem cell, blood transfusion, antimalarial, RAAS inhibitors, anticoagulation, convalescent plasma, immunothrombosis, PD-1 inhibition, immunosuppressants, TNF Inhibition, mRNA translation inhibition |

Tweets that could not be categorized into a domain and subdomain were considered noise. The remaining signal tweets were collected and analyzed for their information density. We analyzed the metadata of each signal tweet to identify those that contained an image(s) and/or a URL linking to an external source. Such tweets were designated as information density signal tweets and were archived in an open-access, searchable, online repository that we created using Google Data Studio. The repository can be accessed at NephTwitterArchive.com. We measured the number of completed searches performed in the archive to gauge provider interest and/or need for COVID-19-related information prior to and after the WHO declared COVID-19 a pandemic (Day 0). A completed search was registered when the provider satisfied four criteria: a) input a query AND b) executed the search AND c) was presented with results AND d) clicked on at least one link to the primary tweet. To encourage use of the Archive, we advertised its existence on Twitter and maintained user anonymity by not collecting user-specific data such as name, location, or provider type.

## Results

During the first 200 days of the pandemic we collected 21071 tweets from the eight physician-established hashtags studied. Only 9051 (43% tweets were considered signal: tweets categorized into both a domain and subdomain. There was a trend towards fewer and fewer signal tweets as the pandemic progressed, with a daily median of 22% (IQR 0-42% (FIGURE 1). The domains with the most tweets were Prevention (2915 tweets) and Therapeutics (3027 tweets) (TABLE 2). Tweets about Prevention were composed at a steady rate from Days 0 to 49 while tweets about Therapeutics peaked on Day 21, approximately seven days before multiple pharmaceutical companies announced vaccine trials (FIGURE 2) (11).

**Table 2.**
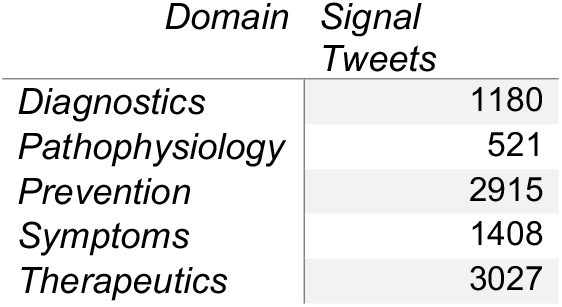

| *Domain* | *Signal*<br>*Tweets* |
| --- | --- |
| *Diagnostics* | 1180 |
| *Pathophysiology* | 521 |
| *Prevention* | 2915 |
| *Symptoms* | 1408 |
| *Therapeutics* | 3027 |

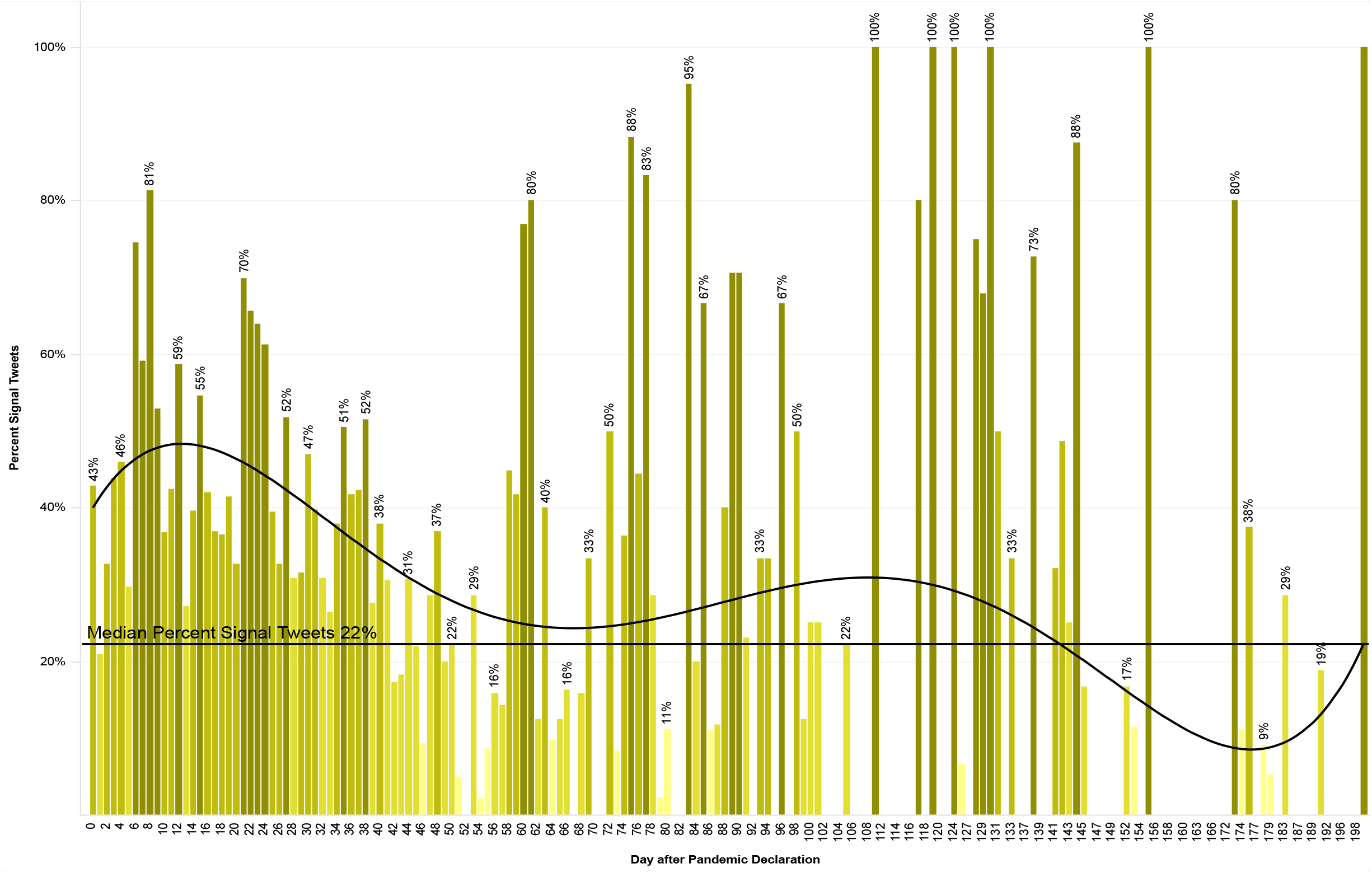

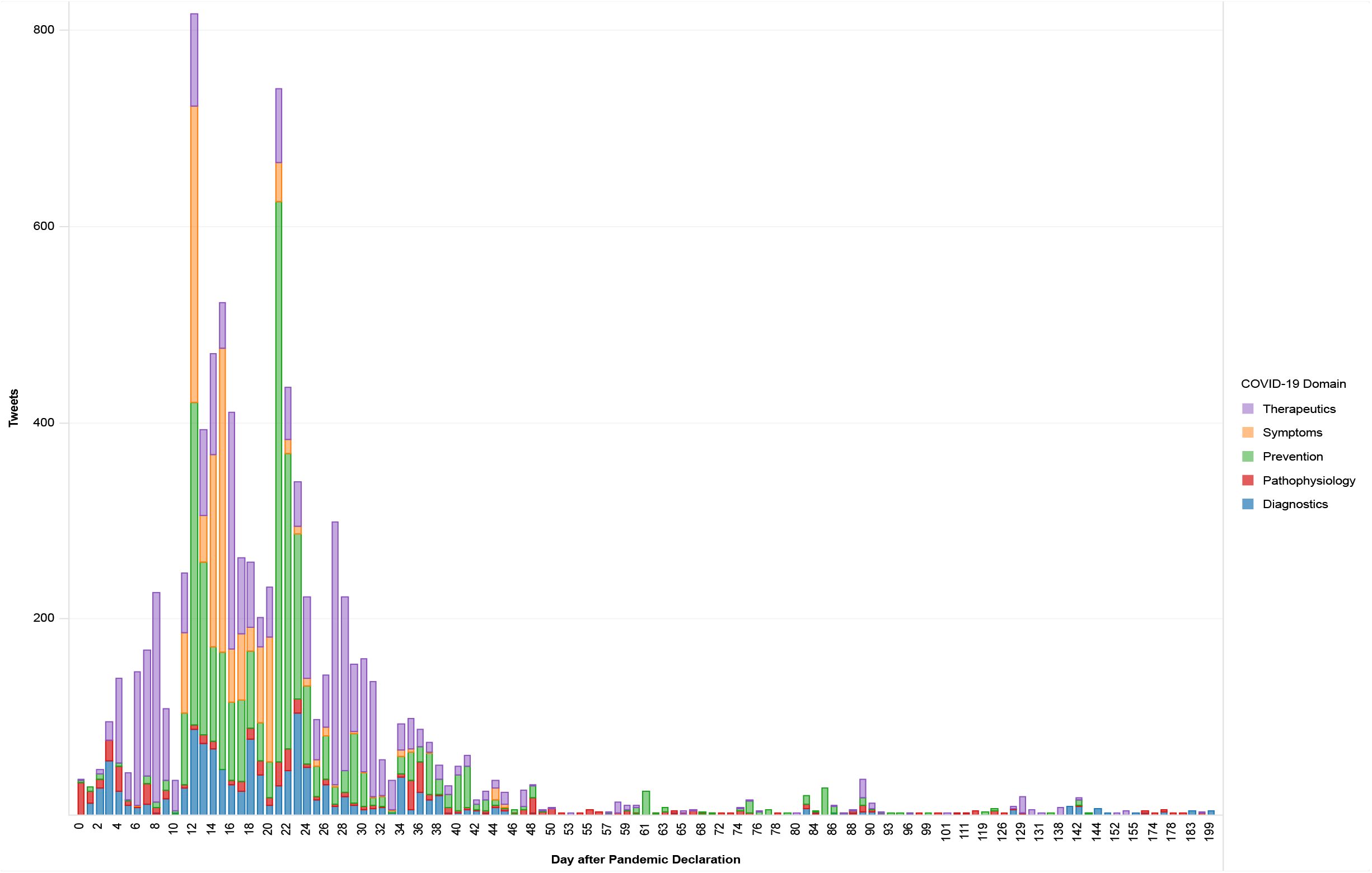

The most popular subdomain in Prevention was PPE (personal protective equipment) (2448 signal tweets). In Therapeutics, Hydroxychloroquine/chloroquine wwo Azithromycin and Mechanical Ventilation were the most popular subdomains (1284 and 1224 signal tweets, respectively) (TABLE 3). Interestingly, neither vaccines nor Remdesivir were heavily discussed in the signal tweets. The Infodemic quieted significantly after Day 50, when the National Institute of Allergy and Infectious Diseases (NIAID) commented about the use of Remdesivir in the Adaptive COVID-19 Treatment Trial (11-13) (TABLE 4).

**Table 3.**
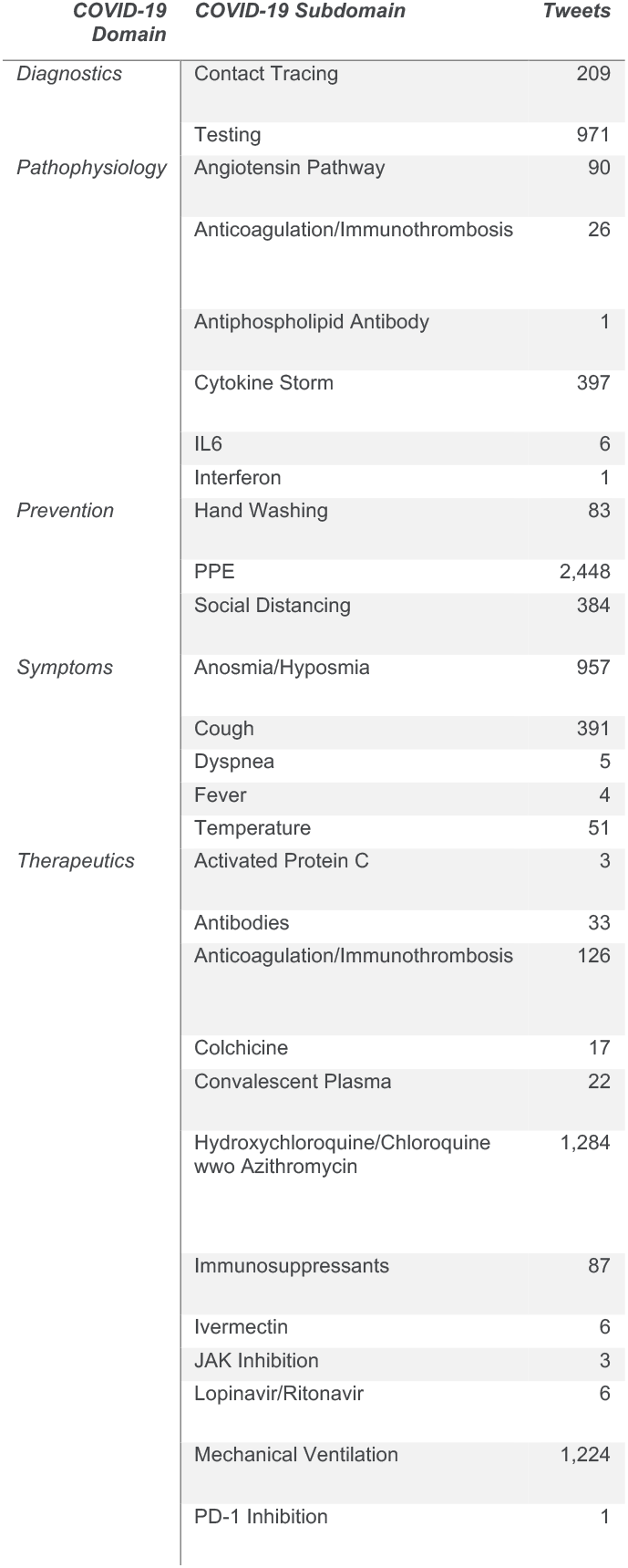
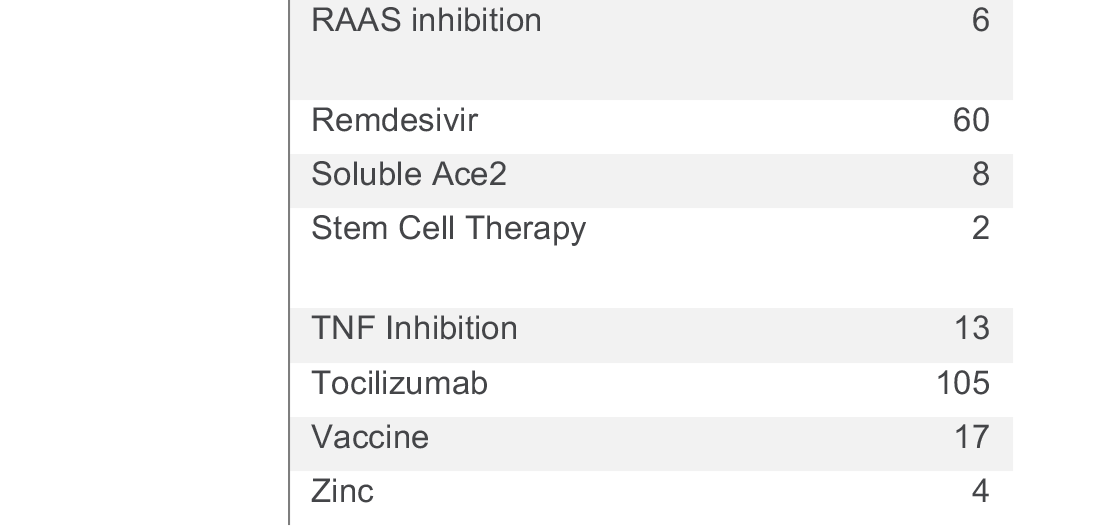

| **COVID-19 Domain** | **COVID-19 Subdomain** | **Tweets** |
| --- | --- | --- |
| *Diagnostics* | Contact Tracing | 209 |
|  | Testing | 971 |
| *Pathophysiology* | Angiotensin Pathway | 90 |
|  | Anticoagulation/Immunothrombosis | 26 |
|  | Antiphospholipid Antibody | 1 |
|  | Cytokine Storm | 397 |
|  | IL6 | 6 |
|  | Interferon | 1 |
| *Prevention* | Hand Washing | 83 |
|  | PPE | 2,448 |
|  | Social Distancing | 384 |
| *Symptoms* | Anosmia/Hyposmia | 957 |
|  | Cough | 391 |
|  | Dyspnea | 5 |
|  | Fever | 4 |
|  | Temperature | 51 |
| *Therapeutics* | Activated Protein C | 3 |
|  | Antibodies | 33 |
|  | Anticoagulation/Immunothrombosis | 126 |
|  | Colchicine | 17 |
|  | Convalescent Plasma | 22 |
|  | Hydroxychloroquine/Chloroquine<br>wwo Azithromycin | 1,284 |
|  | Immunosuppressants | 87 |
|  | Ivermectin | 6 |
|  | JAK Inhibition | 3 |
|  | Lopinavir/Ritonavir | 6 |
|  | Mechanical Ventilation | 1,224 |
|  | PD-1 Inhibition | 1 |

|  |  |
| --- | --- |
| RAAS inhibition | 6 |
| Remdesivir | 60 |
| Soluble Ace2 | 8 |
| Stem Cell Therapy | 2 |
| TNF Inhibition | 13 |
| Tocilizumab | 105 |
| Vaccine | 17 |
| Zinc | 4 |

**Table 4.**

| ***Pandemic Day*** | **Key Event** |
| --- | --- |
| 0 | WHO declares COVID-19 a pandemic |
| 8 | US President suggests Hydroxychloroquine as potential treatment |
| 28 | Multiple pharma companies announce vaccine trials |
| 34 | US removes funding for WHO |
| 36 | VA Healthcare study shows increased death risk with HCQ in COVID-19 |
| 49 | NIAID statement on ACTT-1: remdesivir accelerates recovery |

We evaluated the signal tweets and identified 2996 (33% being the highest quality: containing either a citation to an external source or an image(s) or both. These tweets were archived in NephTwitterArchive.com and available for searches. During the active Infodemic phase (Days 0 to 49), a total of 2021 searches were completed, which was a 26% increase from the same time period before the pandemic was declared (Days −50 to −1). The average daily completed searches was 41.2 in the active Infodemic period; daily completed searches increased to 47.6 during the quiescent period (Days 50-199; 7144 completed searches).

## Discussion

The COVID-19 Infodemic exacerbated an already known and increasingly difficult challenge for those who rely on Twitter to share scientific information: the number of low-quality tweets is overwhelmingly abundant and easily dilutes higher quality tweets (7-8). We coded algorithms to separate signal from noise. By categorizing tweets into domains and subdomains, we were able to reduce the noise by 43%. Subsequently, we identified and archived only 33% of the categorized tweets. By comparing completed searches pre- and post-pandemic (days −50 to −1 and days 0 to +49, respectively), we were able to gauge interest in COVID-19-specific tweets by the Twitter healthcare community. The substantial increase in searches suggests that healthcare providers were looking for this information. Two representative examples show the limitations and strengths of our algorithms. We believe the significant signal activity about Hydroxychloroquine/chloroquine wwo Azithromycin had more to do with a political misunderstanding of its efficacy (or lack thereof) rather than a legitimate scientific controversy about its effectiveness (14). Our algorithms could not distinguish political misunderstanding from legitimate scientific controversy and thus tweets about hydroxychloroquine were classified as signal and some were archived in NephTwitterArchive.com. We were surprised that neither vaccines or the drug Remdesivir were actively discussed. The lack of signal tweets for either subdomain may suggest that not enough scientific information and/or clinical experience was available to generate informative tweets (15). In this case, our algorithms identified these tweets as unsupported by external evidence and eliminated them from the archive.

## Conclusion

The Infodemic indicates that future endeavors must be undertaken to eliminate noise and elevate signal in all aspects of scientific discourse on Twitter (16). Our strategy to eliminate noise and elevate signal has limitations, but in the absence of any algorithm-focused strategy, healthcare providers will be left with the nearly impossible task of manually finding high-quality tweets from amongst a tsunami of noise.

## Data Availability

All data is available and updated at NephTwitterArchive.com

https://NephTwitterArchive.com

## References

1. Zimba O, Radchenko O, Strilchuk L (2020) Social media for research, education and practice in rheumatology. Rheumatol Int 40(2):183–190. https://doi.org/10.1007/s00296-019-04493-4

2. Fischman DL, Mamas MA, Alasnag M, Parwani P, Savage, MP, Desai T. Understanding the Analytics of Twitter in Cardiovascular Medicine. J Am Coll Cardiol Case Rep. 2020. 5:837–839

3. Pemmaraju N, Thompson MA, Mesa RA, Desai T. Anaylsis of the Use and Impact of Twitter During American Society of Clinical Oncology Annual Meetings From 2011 to 2016: Focus on Advanced Metrics and User Trends. Journal of Oncology Practice. 2017. DOI: 10.1200/JOP.2017.021634

4. Desai T. Merging the Traditional with the New: CME-accredited Twitter Journal Clubs. medRxiv. 2020. DOI: 10.1101/2020.04.22.20075606

5. Thamman R, Desai T, Wiener DH, Swaminathan M. ASEchoJC Twitter Journal Club to CME: A Paradigm Shi in Cardiology Education. Journal of the American Society of Echocardiography. 2020; 33(3): A29. DOI: 10/1016/h.echo.2020.01.003

6. Chan AKM, Nickson CP, Rudolph JW, Lee A, Joynt GM. Social media for rapid knowledge dissemination: early experience from the COVID-19 pandemic. Anaesthesia 2020. doi: 10.1111/anae.15057 G. Allen HG, Stanton TR, et al. Social media release increases dissemination of original articles in the clinical pain sciences. PLoS One. 2013;8(7):e68914

7. Marr B. Coronavirus Fake News: How Facebook, Twitter, and Instagram Are Tackling The Problem. Forbes 2020. Mar 27. https://www.forbes.com/sites/forbes-personal-shopper/2020/10/26/best-humidifiers-2020/40b0edeb7f81

8. Desai T. Physician-initiated Research in Social Media. An Area That Can No Longer Be Ignored. ATS Scholar. 2019. DOI: 10.34197/ats-scholar.2019-0026ED

9. World Health Organization. Novel Coronavirus(2019-nCoV). Situation Report-13. 2 February 2020. https://www.who.int/docs/default-source/coronaviruse/situation-reports/20200202-sitrep-13-ncov-v3.pdf

10. Lamsal R. Coronavirus (COVID-19) Tweets Dataset. IEEE Dataport 2020. Dx.doi.org/10.21227/781w-ef42

11. Taylor DB. A Timeline of the Coronavirus Pandemic. The New York Times. 2020. https://nyti.ms/3afns4U

12. NIH Clinical Trial Shows Remdesivir Accelerates Recovery from Advanced COVID-19. 2020. https://www.niaid.nih.gov/news-events/nih-clinical-trial-shows-remdesivir-accelerates-recovery-advanced-covid-19

13. Beigel JH, Tomashek KM, Dodd LE, Mehta AK, et al. Remdesivir for the Treatment of Covid-19 – Final Report. NEJM. 2020. DOI: 10.1056/NEJMoa2007764

14. Sebastian E. Sattui, Jean W. Liew, Elizabeth R. Graef, Ariella Coler-Reilly, Francis Berenbaum, Ali Duarte-Garcia, Carly Harrison, Maximilian F. Konig, Peter Korsten, Michael S. Putman, Philip C. Robinson, Emily Sirotich, Manuel F. Ugarte-Gil, Kate Webb, Kristen J. Young, Alfred H.J. Kim Jerey A. Sparks (2020) Swinging the pendulum: lessons learned from public discourse concerning hydroxychloroquine and COVID-19, Expert Review of Clinical Immunology, 16:7, 659–666, DOI: 10.1080/1744666X.2020.1792778

15. Liu M, Caputi TL, Dredze M, Kesselheim AS, Ayers JW. Internet Searches for Unproven COVID-19 Therapies in the United States. JAMA Internal Medicine. 2020;180(8):1116–1118. doi:10.1001/jamainternmed.202

16. Pollett S, Rivers C (2020) Social media and the new world of scientific communication during the COVID19 pandemic. Clin Infect Dis. https://doi.org/10.1093/cid/ciaa553

